# ChatGPT for Clinical Vignette Generation, Revision, and Evaluation

**DOI:** 10.1101/2023.02.04.23285478

**Authors:** James R. A. Benoit

## Abstract

**Objective:** To determine the capabilities of ChatGPT for rapidly generating, rewriting, and evaluating (via diagnostic and triage accuracy) sets of clinical vignettes.

**Design:** We explored the capabilities of ChatGPT for generating and rewriting vignettes. First, we gave it natural language prompts to generate 10 new sets of 10 vignettes, each set for a different common childhood illness. Next, we had it generate 10 sets of 10 vignettes given a set of symptoms from which to draw. We then had it rewrite 15 existing pediatric vignettes at different levels of health literacy. Fourth, we asked it to generate 10 vignettes written as a parent, and rewrite these vignettes as a physician, then at a grade 8 reading level, before rewriting them from the original parent’s perspective. Finally, we evaluated ChatGPT for diagnosis and triage for 45 clinical vignettes previously used for evaluating symptom checkers.

**Setting and participants:** ChatGPT, a publicly available, free chatbot.

**Main outcome measures:** Our main outcomes for de novo vignette generation were whether ChatGPT followed vignette creation instructions consistently, correctly, and listed reasonable symptoms for the disease being described. For generating vignettes from pre-existing symptom sets, we examined whether the symptom sets were used without introducing extra symptoms. Our main outcome for rewriting existing standardized vignettes to match patient demographics, and rewriting vignettes between styles, was whether symptoms were dropped or added outside the original vignette. Finally, our main outcomes examining diagnostic and triage accuracy on 45 standardized patient vignettes were whether the correct diagnosis was listed first, and if the correct triage recommendation was made.

**Results:** ChatGPT was able to quickly produce varied contexts and symptom profiles when writing vignettes based on an illness name, but overused some core disease symptoms. It was able to use given symptom lists as the basis for vignettes consistently, adding one additional (though appropriate) symptom from outside the list for one disease. Pediatric vignettes rewritten at different levels of health literacy showed more complex symptoms being dropped when writing at low health literacy in 87.5% of cases. While writing at high health literacy, it added a diagnosis to 80% of vignettes (91.7% correctly diagnosed). Symptoms were retained in 90% of cases when rewriting vignettes between viewpoints. When presented with 45 vignettes, ChatGPT identified illnesses with 75.6% (95% CI, 62.6% to 88.5%) first-pass diagnostic accuracy and 57.8% (95% CI, 42.9% to 72.7%) triage accuracy. Its use does require monitoring and has caveats, which we discuss.

**Conclusions:** ChatGPT was capable, with caveats and appropriate review, of generating, rewriting, and evaluating clinical vignettes.

## Introduction

Clinical vignettes are written case reports and simulations used in health education and research. Vignettes are designed to elicit readers’ beliefs,^1^ and aim to elicit similar responses in hypothetical situations as would be shown in the real world.^2^ However, for such a widespread teaching and practice tool, vignette development or validation is a relative unknown, and there has been debate around the applicability of vignettes to actual decision-making.^3^ Development of these health education items is also expensive, with a similarly constructed item, multiple choice test questions, estimated at a cost of $1500-2500 each.^4^ Previous studies have proposed systematic frameworks for developing vignettes for health professionals,^2^ methods for evaluating vignette content validity,^5^ and integrating co-design into the vignette creation process.^6^ A recent review that aimed to synthesize vignette methodology in qualitative healthcare research suggested that this methods-focused work was necessary, arguing that the variability between development methods demonstrated the need for a more rigorous, transparent vignette development process.^7^ However, there is little work into generating large volumes of clinical vignettes in an automated way. While variations of a technique called Automated Item Generation (AIG) (a template-based, expert-informed process)^4^ have been used in the past, we were unable to find examples in the literature of using chatbots or conversational AI for this purpose. This study explores the use of a modern conversational agent (aka chatbot), specifically ChatGPT, for generating and evaluating clinical vignettes.

Chatbots are computer programs that simulate human-like conversations, and have become increasingly popular with advancements in natural language processing (NLP, a branch of Artificial Intelligence (AI) focusing on linguistic interactions between humans and computers).^8^ Chatbots are used in a wide range of applications such as marketing and education,^9^ and in health, including online medical consultation (Babylon Health), Florence the “personal nurse” reminding patients to take their medications, and diagnostic agents such as Ada Health, a virtual symptom checker.^10^ Chatbots can be based on a number of theoretical foundations. The first chatbots were based on templates: conversations were created by pattern-matching user inputs to predefined outputs. As technology evolved, chatbots were created to handle large amounts of data (corpus-based chatbots), then respond more flexibly by addressing the intent of user queries (intent-based chatbots), generate their own dialogue rather than responding based on pattern-matching (Recurrent Neural Network (RNN)-based chatbots), and using the context in which queries were made to inform their responses (Reinforcement Learning (RL)-based chatbots).^11^ From these approaches, a hybrid model was developed that asks more than one chatbot to contribute to its response, using a ranking system to generate a combined output. This approach has led to the recent development of large language models: a group of machine learned algorithms trained on massive amounts of data that are aimed at generating human-like text (e.g. translation, summarization, and conversation).^12^ These chatbots belong to a larger class of AI called generative models. Generative models are being used in research to write papers (e.g. Galactica), code (e.g. Codex),^13^ and algorithms (e.g. AlphaTensor). These models are now capable of translating between modes of communication, including text-to-audio (e.g. AudioLM),^14^ text-to-image (e.g. DALLE-2),^15^ text-to-3D image (e.g. Dreamfusion),^16^ and text-to-video (Phenaki).^17^ The opposite way is possible as well, with models such as Flamingo,^18^ enabling image-to-text generation.^19^

Chatbots have struggled with several issues, including the ability to understand colloquial language, their flexibility in providing solutions to new problems, and trained-in bias (negative real-world effects caused by the data on which the chatbot was trained). ChatGPT is a chatbot running a 175 billion-parameter language model, made by combining a large language model called GPT-3 (Generative Pre-trained Transformer 3) and fine-tuning its responses using Reinforcement Learning with Human Feedback (RLHF).^20^ In the first months since its release on November 30 2022, ChatGPT’s ability to respond flexibly to ever-increasing demands (from simple queries to writing full scientific papers) has generated enough interest for more than 1350 research articles on the topic, indexed by Google Scholar. We began exploring ChatGPT because of its potential to generate large, standardized, high-quality databases of clinical vignettes relevant to our work with common childhood illnesses.^21^ In this study, we will examine how well ChatGPT can generate clinical vignettes from simple prompts and predefined symptom sets, rewrite vignettes to match the communication styles of different audiences, and evaluate existing vignettes using the same language model.

## Methods

We used ChatGPT (Jan 9 version, Free Research Preview),^22^ as the basis of five experiments to explore its capabilities in vignette generation from a simple text prompt, identify diseases and generate vignettes from predefined symptom sets, rewrite vignettes with different levels of medical vocabulary, and rewrite given vignettes in different communication styles. We also evaluated its capacity for diagnosis and triage of an existing set of 45 standardized vignettes.^21^

### Experiment 1

First, we generated a synthetic set of pediatric clinical vignettes using ChatGPT. We created 10 pediatric clinical vignettes for each of 10 common childhood diseases, each using a prompt given below. The set of diseases is listed in Table 1.

**Table 1.**
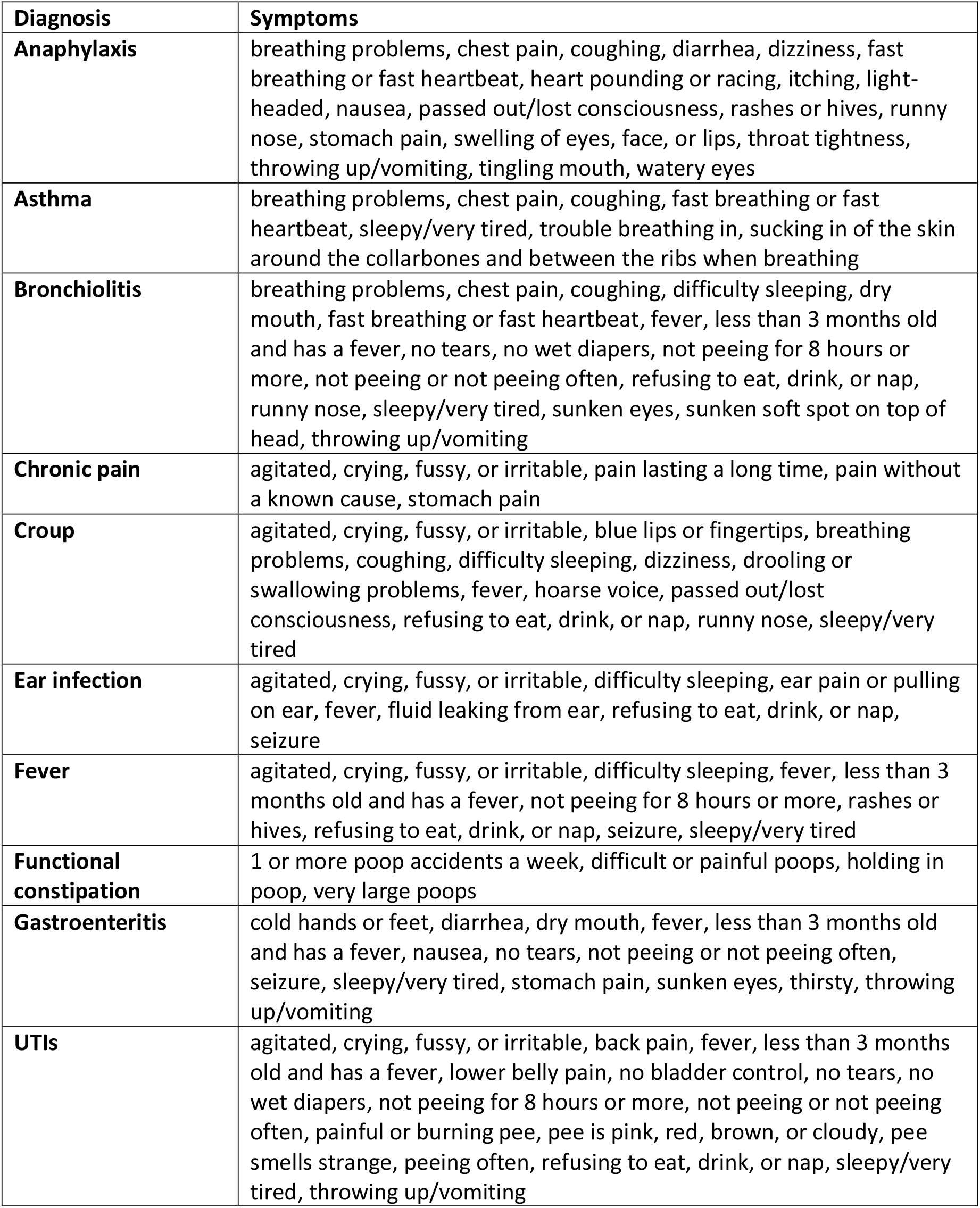
List of symptoms used to prompt ChatGPT, and associated illness.

#### Prompt

Write ten different clinical vignettes in the first person from the perspective of the child’s mother or father or guardian speaking. Each vignette should contain one to all of the lay symptoms of a child who has [disease]. Do not use the term [disease], but make sure the combination of symptoms you use points to it. Do not use symptoms from the previous vignette.

We then input the set of prompts for each disease into ChatGPT to summarize symptom counts.

### Experiment 2

We had ChatGPT write pediatric clinical vignettes based on a predefined symptom list. Symptoms were initially entered as a list for each disease, with no additional prompt given, and ChatGPT was allowed to respond. ChatGPT was then prompted to write 10 vignettes based on each symptom list. We used a list of symptoms drawn from existing knowledge translation tools (KT tools), which were developed using best-available evidence and co-developed with parents, healthcare professionals, and researchers.^23-32^ This list of symptoms was developed for use in a symptom checker for differentiating between diseases, and is not intended to provide a complete list of symptoms describing each disease.

#### Initial prompt

List of symptoms for one disease, as shown in Table 1.

#### Follow-up Prompt

Using the list of symptoms above, generate 10 pediatric clinical vignettes about [disease]. Do not use the term [disease].

### Experiment 3

We explored how well ChatGPT could apply different levels of medical vocabulary to an existing vignette. We asked ChatGPT to explain its understanding of health literacy before asking it to rewrite a prompt, to gain understanding of its knowledge base in this domain. We asked it to rewrite the 15 pediatric clinical vignettes from Semigran et al. (2015), as written in the first person by parents at low, average, and high health literacy.

#### Comprehension prompt

What is health literacy?

#### Prompt 1

Rewrite the following clinical vignette, in first person, as a child’s parent, in each of the following styles: the parent has low, average, or high health literacy. Identify the style in which you wrote each prompt. Vignette: [Semigran vignette]

### Experiment 4

Our fourth experiment examined how well ChatGPT could translate one description of disease between very different communication styles, while maintaining the same information content. We asked it to write ten vignettes about anaphylaxis from a parent’s perspective, using a given list of symptoms. Then, ChatGPT was prompted to rewrite these as standard clinical vignettes using medical vocabulary. We then asked it to re-write these vignettes using medical vocabulary at a grade-8 reading level. Finally, we asked it to re-write these back to their original form: in the first person as a worried parent.

#### Initial Prompt

Write pediatric clinical vignettes for [disease] from the first-person perspective of a worried parent, using partial sets of the following set of symptoms: [symptom list for disease].

#### Follow-up prompt

Re-write as a clinical prompt from the perspective of a clinician.

#### Follow-up prompt 2

Re-write at a grade-8 reading level.

#### Follow-up prompt 3

Re-write in the first person as a worried parent.

### Experiment 5

Finally, we examined how well ChatGPT could match diagnoses from existing clinical vignettes. We asked it to diagnose and triage into the three premade categories (Emergent care required/non-emergent care reasonable/self-care reasonable) a set of 45 simplified standardized patient vignettes for assessing symptom checkers.^21^ We also tested diagnosis of the full prompts to discover any differences in primary diagnosis caused by how the prompt was presented to ChatGPT.

#### Prompt

Diagnose [Semigran (2015) simplified vignette].

Follow-up prompt: Consider the following simplified clinical vignettes. For each, triage it into one of three categories: emergent care required (for example, pulmonary embolism), non-emergent care reasonable (for example, otitis media), and self care reasonable (for example, viral upper respiratory tract infection). First vignette [Semigran (2015) simplified vignette].

#### Prompt 2

Diagnose [Semigran (2015) full vignette]

## Results

### Experiment 1

We began by asking ChatGPT to generate clinical vignettes in lay language and add variation by avoiding symptoms in the previous vignette, and avoiding the name of the disease, but still providing a clear description of symptoms that would be diagnosed as that disease. This was generated as part of a varied dataset of vignettes to be used for symptom checker evaluation. The vignettes produced were short, to-the-point, and contained basic symptom information, and were all written in the first person. ChatGPT did not propose symptoms that would be unexpected in the 10 diseases evaluated. However, it did use the term, “fever,” in 30% of Fever vignettes, and the same symptom being used in subsequent vignettes occurred 20.5% more than parameters specified. Further testing of this phenomenon revealed that ChatGPT was unwilling to write a set of 10 vignettes without using the same symptoms in adjacent prompts, if it considered the symptom to be key to the disease. Instead, it switched other rules, using different symptom combinations or contexts in which the symptom were presented. According to ChatGPT, “If I have to write ten clinical vignettes, each with different symptoms, I would still make sure to include the key symptoms of the disease in each vignette. Although I need to change the combinations or context of the symptoms, it is important to accurately reflect the most important signs of the disease in each vignette.” Figure 1 shows the presence of all symptoms occurring six or more times (and therefore in at least one subsequent set of vignettes).

**Figure 1.**
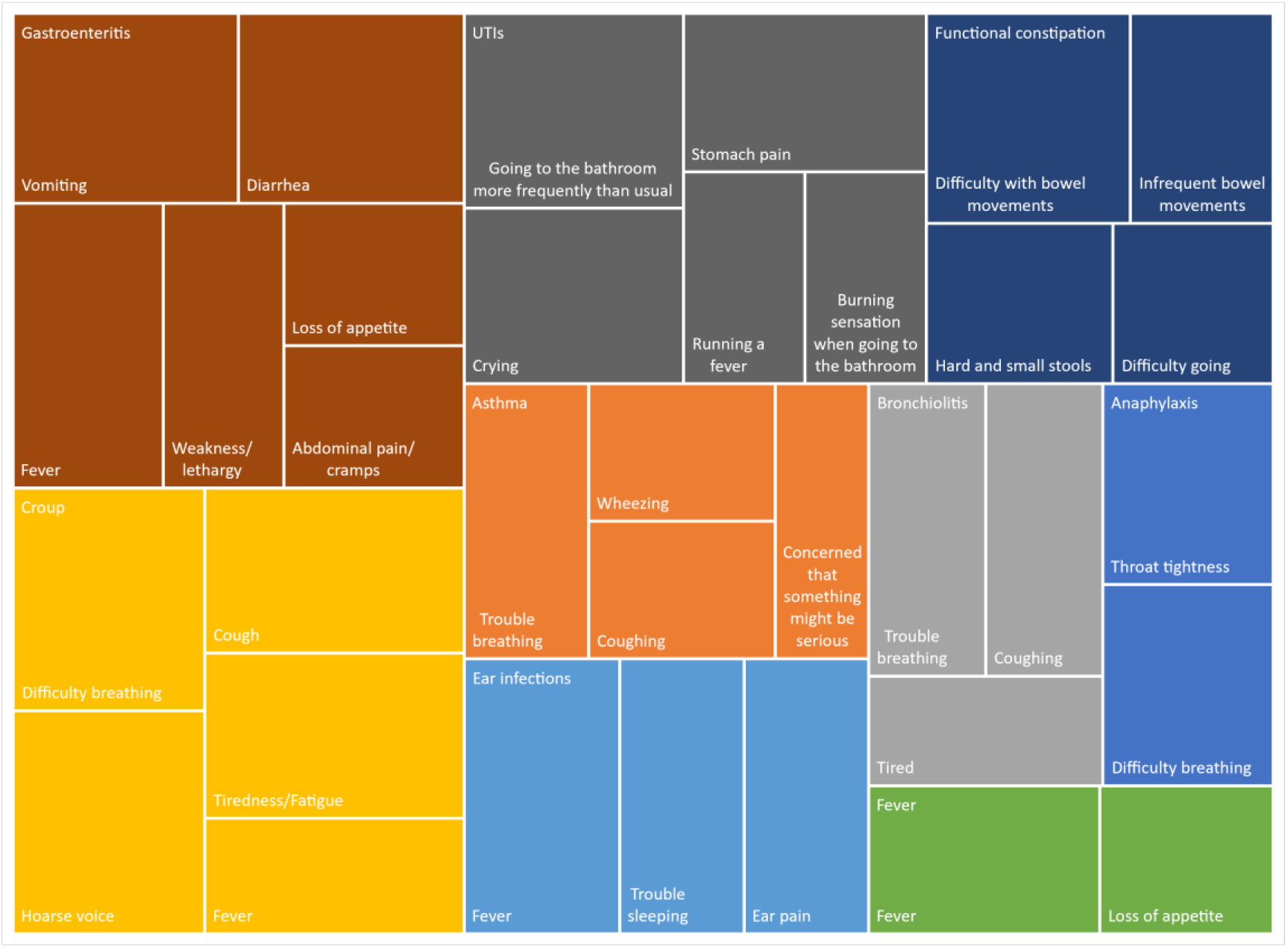
Relative frequency of symptoms occurring in 60% or more of vignettes for that illness.

**Table 2.**
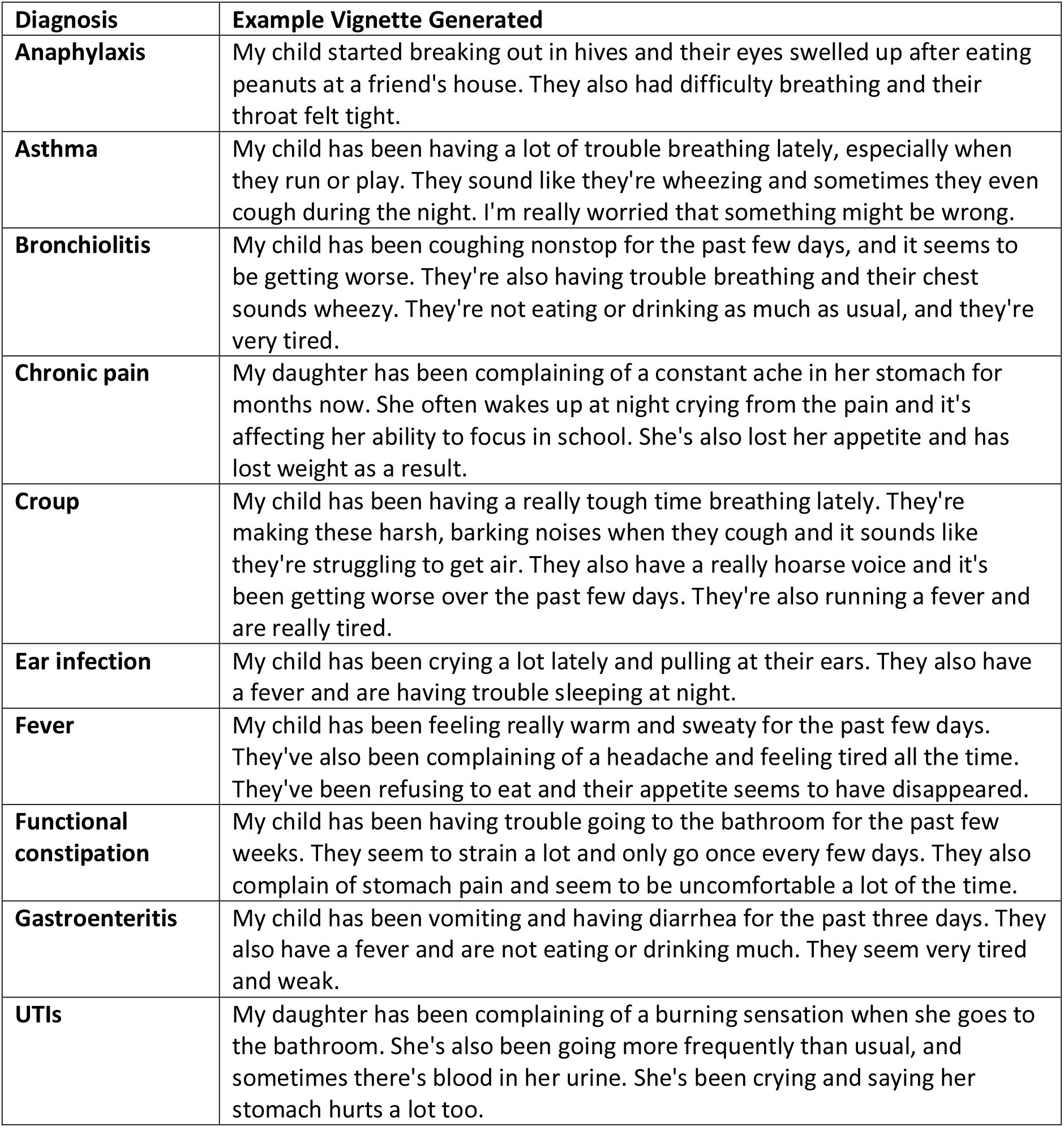
Representative short vignettes written to describe each of 10 common children’s illnesses.

### Experiment 2

ChatGPT wrote vignettes using predefined symptom lists as short case reports. It did not include symptoms outside the list specified in 90% of diseases, except as context for the illness (e.g. difficulty playing resulting from coughing or difficulty breathing). However, there was an exception for functional constipation, where all vignettes had a variation of abdominal pain added.

**Table 3.**
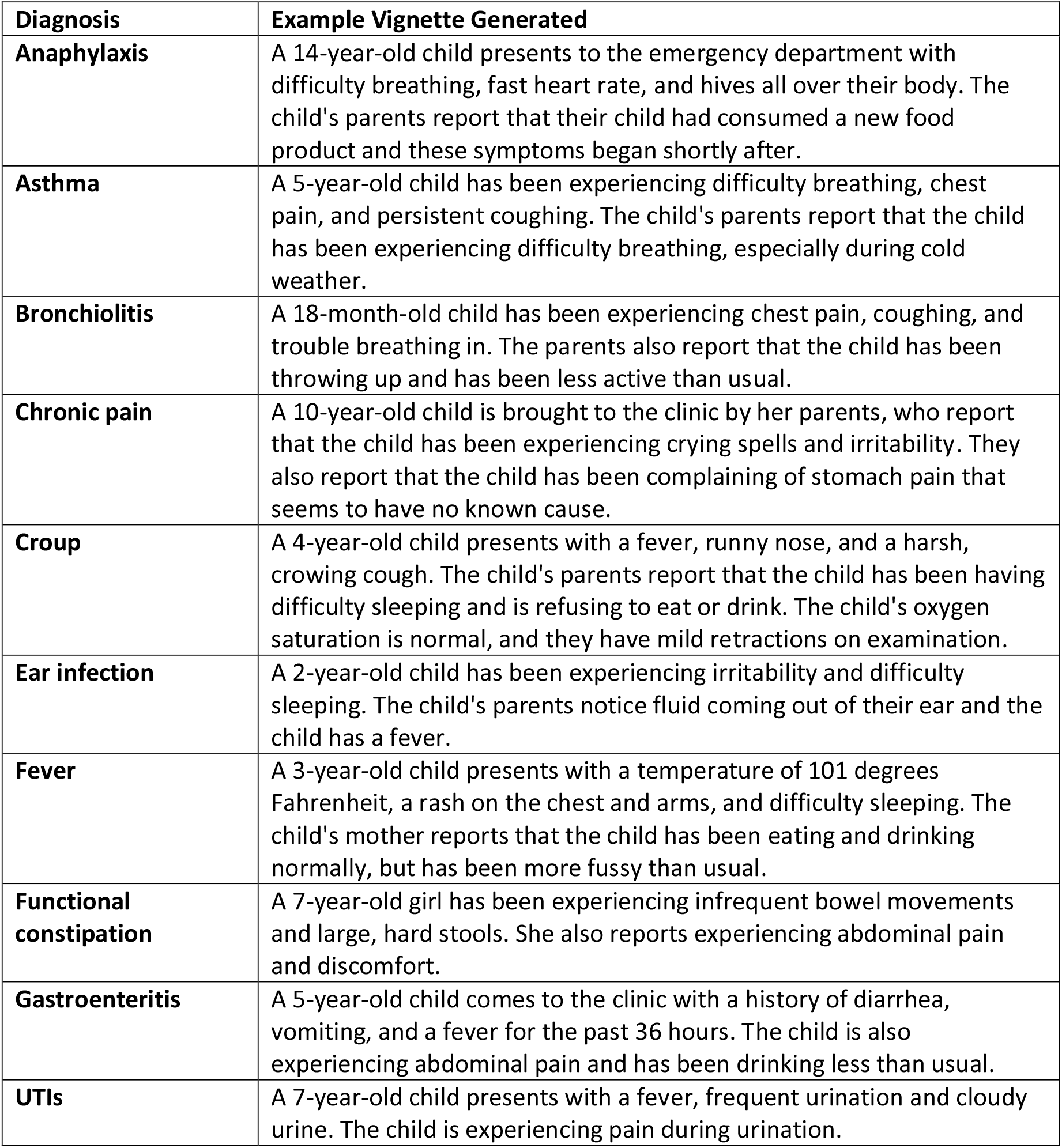
Vignettes based on provided symptom profile.

| Diagnosis | **Example Vignette Generated** |
|---|---|
| **Anaphylaxis** | A 14-year-old child presents to the emergency department with difficulty breathing, fast heart rate, and hives all over their body. The child's parents report that their child had consumed a new food product and these symptoms began shortly after. |
| **Asthma** | A 5-year-old child has been experiencing difficulty breathing, chest pain, and persistent coughing. The child's parents report that the child has been experiencing difficulty breathing, especially during cold weather. |
| **Bronchiolitis** | A 18-month-old child has been experiencing chest pain, coughing, and trouble breathing in. The parents also report that the child has been throwing up and has been less active than usual. |
| **Chronic pain** | A 10-year-old child is brought to the clinic by her parents, who report that the child has been experiencing crying spells and irritability. They also report that the child has been complaining of stomach pain that seems to have no known cause. |
| **Croup** | A 4-year-old child presents with a fever, runny nose, and a harsh, crowing cough. The child's parents report that the child has been having difficulty sleeping and is refusing to eat or drink. The child's oxygen saturation is normal, and they have mild retractions on examination. |
| **Ear infection** | A 2-year-old child has been experiencing irritability and difficulty sleeping. The child's parents notice fluid coming out of their ear and the child has a fever. |
| **Fever** | A 3-year-old child presents with a temperature of 101 degrees Fahrenheit, a rash on the chest and arms, and difficulty sleeping. The child's mother reports that the child has been eating and drinking normally, but has been more fussy than usual. |
| **Functional constipation** | A 7-year-old girl has been experiencing infrequent bowel movements and large, hard stools. She also reports experiencing abdominal pain and discomfort. |
| **Gastroenteritis** | A 5-year-old child comes to the clinic with a history of diarrhea, vomiting, and a fever for the past 36 hours. The child is also experiencing abdominal pain and has been drinking less than usual. |
| **UTIs** | A 7-year-old child presents with a fever, frequent urination and cloudy urine. The child is experiencing pain during urination. |

### Experiment 3

#### Comprehension prompt response

Health literacy is the ability of individuals to access, understand, and use health information and services to make informed decisions about their health. This includes being able to read and understand medical instructions, navigate the healthcare system, and communicate effectively with healthcare providers. Low health literacy can lead to poor health outcomes, as individuals may be unable to access the information and services they need to manage their health effectively.

**Table 4.**
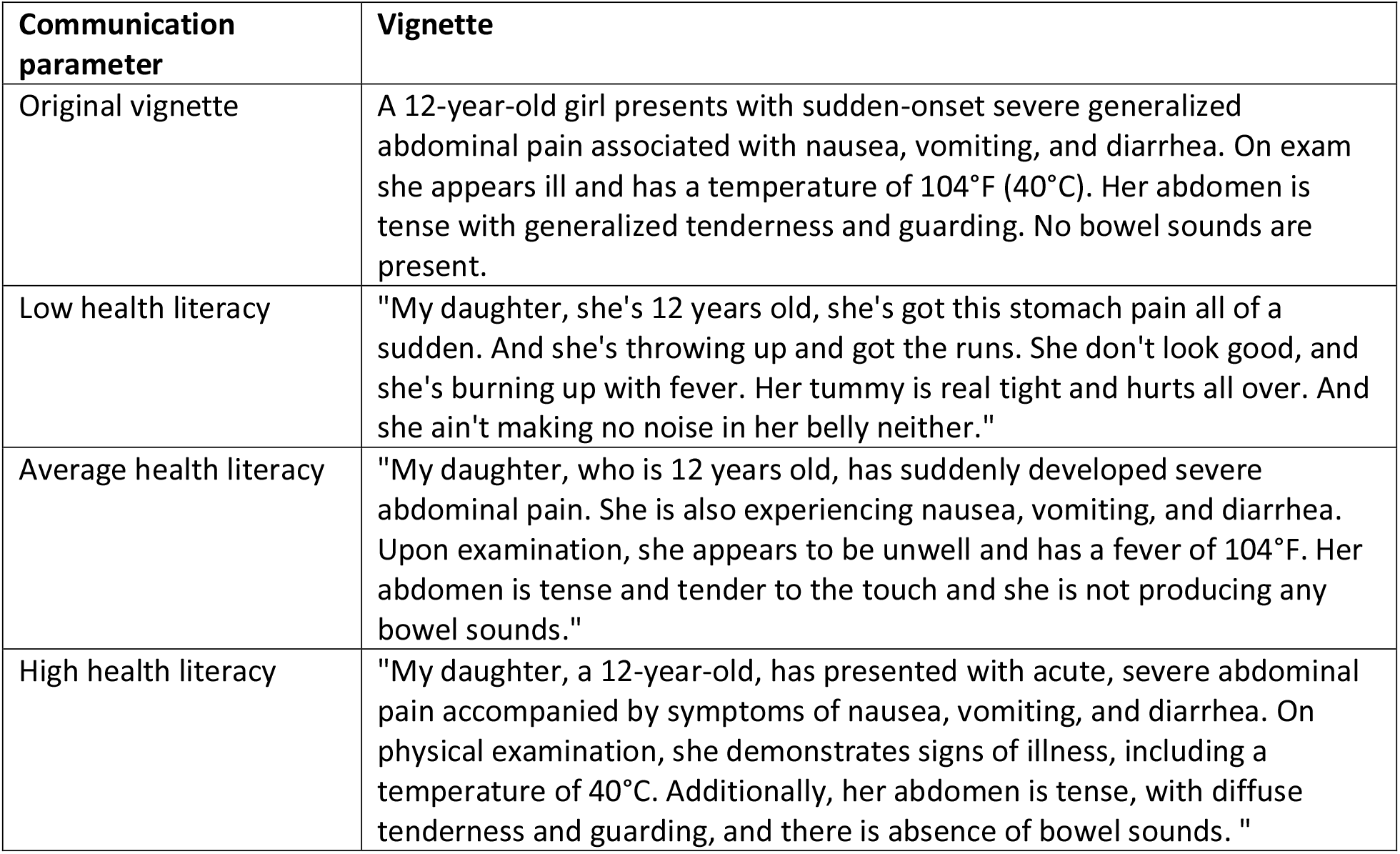
Example of different communication style outputs by ChatGPT

Differences were seen between the vocabulary used to describe symptoms at low and high health literacy (e.g., “… her tummy is real tight and hurts all over.” vs. “…her abdomen is tense, with diffuse tenderness and guarding…”), and grammar. In addition, the detail in which symptoms (e.g. temperature) were presented increased with health literacy. ChatGPT showed three behaviours of note on this task. It chose not to add more complex symptoms to the low health literacy vignettes (e.g., “Enlarged cervical lymph nodes, exudative pharyngitis with soft palate petechiae and faint erythematous macular rash on the trunk and arms are found”), in 87.5% (n=8) of vignettes containing extended symptom descriptions. There were two instances where it equated nausea and vomiting, and one instance where it lumped a lack of specific symptoms together as, “…nothing else is wrong with him.” Finally, it added a diagnosis to the high health literacy category in 80% (n=12) of pediatric vignettes (and was correct in 91.7% of diagnoses added), with one labeled as incorrect for lacking specificity when identifying the given diagnosis of salmonella as, “food-borne illness.”

It should also be noted that the wording of the low health literacy vignette can be re-written with fewer colloquialisms, and the same symptom names, e.g.:

“My daughter, she’s 12 years old, has developed stomach pain all of a sudden. She’s also throwing up and got the runs. She doesn’t look well and has a fever. Her tummy is really tight and hurts all over. And she’s not making any noise in her belly either.”

### Experiment 4

ChatGPT was capable of translating vignettes between parent, physician, and grade 8 writing styles, and translate this output back to being written as a parent. 90% of vignettes retained all symptoms and content when re-written as a parent, although 30% of vignettes made breathing-related symptoms less specific (i.e. “breathing fast,” or “wheezing,” became “trouble breathing”), one relabeled “light-headed” as “dizzy,” and 1/10 removed rash (first calling it pruritis in the physician-emulating vignette, and removing it between the physician and grade 8 reading level vignettes).

**Table 5.** Re-writing vignettes in different styles.

| **Vignette from parent’s perspective** | **Rewritten parent vignette as a clinical vignette using medical vocabulary** | **Rewritten clinical vignette as appropriate to a grade-8 reading level** | **Rewritten clinical vignette back in parent’s perspective.** |
|---|---|---|---|
| My child started coughing and had trouble breathing after eating a peanut butter sandwich. Their face also swelled up and they couldn't stop scratching their skin. | A 4-year-old child presented with symptoms of urticaria and dyspnea following the ingestion of peanut butter. The patient also exhibited angioedema and pruritus. | A child who is 4 years old had trouble breathing and a rash all over their body after eating a sandwich with peanut butter. They also had swollen lips and eyes and couldn't stop scratching their skin. | I was so worried when my 4-year-old child had trouble breathing and a rash all over their body after eating a sandwich with peanut butter. Their lips and eyes were swollen and they couldn't stop scratching their skin. |
| My baby suddenly broke out in hives and started throwing up. Their breathing became very fast and they passed out for a few seconds. | A 6-month-old infant was brought to the clinic with symptoms of vomiting and urticaria. The patient also developed tachypnea and lost consciousness for a brief period. | A baby got very sick and threw up after breaking out in hives. They also had trouble breathing and passed out for a bit. | I was really scared when my baby got very sick and threw up after breaking out in hives. They also had trouble breathing and passed out for a bit. |
| My toddler's throat got tight and they started wheezing after getting a bee sting. They also had swelling around their eyes and lips. | A 2-year-old toddler presented with symptoms of anaphylactoid reaction including laryngeal edema and wheezing following a bee sting. The patient also exhibited periorbital and lip edema. | A toddler's throat felt tight and they had trouble breathing after getting stung by a bee. They also had swollen eyes and lips. | I was so worried when my toddler's throat felt tight and they had trouble breathing after getting stung by a bee. Their eyes and lips were swollen too. |

### Experiment 5

ChatGPT made a correct diagnosis on the first try, on 71.1% of the simplified clinical vignettes, and correctly triaged 57.8% (95% CI, 42.9%-72.7%). A summary of ChatGPT triage recommendations based on the simplified vignettes is presented in Table 6.

**Table 6.** Summary of triage recommendations based on simplified vignettes.

|  | Emergent care required | Non-emergent care reasonable | Self care reasonable |
|---|---|---|---|
| **Emergent care cases** | **12/15 (80%)** | 2/15 (13%) | 0/15 (0%) |
| **Non-emergent care cases** | 3/15 (20%) | **13/15 (87%)** | 14/15 (93%) |
| **Home care cases** | 0/15 (0%) | 0/15 (0%) | **1/15 (7%)** |

It was able to make a correct diagnosis on the first try, on 75.6% (95% CI, 62.6%-88.5%) of the full vignettes presented. Of the 24.4% of cases not correctly identified on the first try, 45.5% (n=5) contained a response with the diagnosis partially or fully present (e.g., diagnosing bacterial pharyngitis when the type of pharyngitis was unknown), or the disease was correct but the cause not specific enough (e.g. diagnosing food poisoning but not inferring Salmonella). In one case, no diagnosis was given, but triage instructions were provided based on the presence of a worrying symptom (dehydration: according to ChatGPT, “…The fact that Jack immediately vomits up the juice after drinking it suggests that he may not be able to keep fluids down. If this is the case, it is important to seek medical attention as soon as possible to prevent dehydration…”).

### Experimental Caveats

There are three caveats we noted while evaluating the capabilities of ChatGPT with clinical vignettes:

1. Scientific papers and links looked real, but the papers themselves did not appear to exist in searches, and no archived pages of given web links were found (e.g. https://www.ama-assn.org/physician-finder) on the Wayback machine. For example, one paper reference given was, “Assessing symptom checkers using simplified vignettes: a systematic review.” PMID: 29134795.” However, PMID 29134795 is a reference for, “Robust and Recyclable Substrate Template with an Ultrathin Nanoporous Counter Electrode for Organic-Hole-Conductor-Free Monolithic Perovskite Solar Cells.”^33^ The paper title was not found on Google Scholar.
2. ChatGPT results must be monitored carefully to ensure the queries used do not result in unexpected behaviour. ChatGPT will change behaviour within a set, and it is important that its outputs are checked carefully.
3. Chat history matters, and ChatGPT will remember and make use of previous conversations-this is important when providing definitions of disease, or symptom sets, as it will remember and apply these parameters to future interactions, until a new conversation is started.

## Discussion

Clinical vignettes are a useful tool for health education,^34^ assessment of healthcare professionals,^35; 36^ healthcare research,^37^ and symptom checker evaluation.^38^ However, little is known about how clinical vignettes for health professionals are produced.^2^ In 2022, large sets of vignettes were still being developed by hand, e.g. for use in evaluating symptom checkers,^39^ and evaluated by humans in order to train an AI. Automated vignette generation has been attempted, e.g. in pharmacy education via breaking vignettes down into qualitative elements,^40^ but lacked the flexibility required to apply the process across healthcare fields.

The primary advantages of using ChatGPT for vignette generation were consistency of writing, rapidity of generating varied results, and flexibility in switching between styles. ChatGPT has been shown to be capable of generating plagiarism-free scientific abstracts,^41^ and the same model being used to generate de novo vignettes mean their use will not infringe on other authors. However, its ability to adapt existing vignettes does raise questions of where ownership begins and ends, and at what point the content being adapted into a different context constitutes a new work. These early results are corroborated by ChatGPT being asked to carry out similar health tasks, such as writing radiological reports: ChatGPT was competent in most cases, but made non-trivial errors that suggested review of its outputs is still necessary.^42^ Further testing will serve to define the boundaries of ChatGPT’s ability and create the evidence necessary to better quantify its abilities.

ChatGPT’s 71.1% first-pass diagnostic performance on 45 simplified vignettes, without any specialized training, is more than double the previously reported 34% average symptom checker performance on the same set of vignettes, and its 58% triage accuracy is comparable to the 57% average across symptom checkers.^21^ Its performance listing the correct diagnosis first on 45 vignettes at 75.6%, now performs similarly to physicians’ 72.1% on the same set of 45 vignettes, although more testing must be done on a larger set of vignettes to establish a more precise measure.^43^ Interestingly, these results show there is a benefit of providing context to ChatGPT’s diagnostic accuracy, raising questions around the appropriateness of using symptom-only diagnosis in future symptom checkers. These early results are corroborated by ChatGPT offering near-passing performance on all sections of the USMLE (United States Medical Licensing Exam).^44^ However, providing appropriate triage advice of these models can and should still be improved. Leveraging data from triage nurse decision-making would be a powerful source of information for training future diagnostic agents in this regard, as this would provide large volumes of data for determining the level of care required for common Emergency Department (ED) complaints. However, the lack of progress on triage performance from over-recommending care also plagues ChatGPT.^45; 46^ ChatGPT averaged 83% accurate triage advice on cases requiring emergent- or non-emergent care, but only 7% accuracy on cases treatable at home, instead recommending non-emergent care in nearly all cases. This overly cautious approach has implications for health system use, if patients’ increasing use of these tools impacts the number of unnecessary ED visits.

When considering how to write queries to generate patient vignettes, the following six communication concepts emerged from conversations with ChatGPT as central to the information it needed to write clinical vignettes, and how to bridge the gap between human communication concepts and its information needs.

1. Perspective-the voice delivering the vignette (e.g. healthcare provider, parent, child)
2. Audience- to whom is the vignette being delivered? (e.g., health professionals, parenting classes)
3. Content- what content must the vignette contain? (e.g., symptoms or symptom sets)
4. Context- how will the vignette will be used? (e.g., for research, for teaching)
5. Structure- how should the narrative flow (e.g., an outline to follow)
6. Tone- emotional perspective of the response (e.g., empathetic, worried)

e.g. “A series of ten clinical vignettes to be used for teaching undergraduate medical classes, written as an empathetic physician, containing the most common symptoms of COVID.”

This work has potential application in medical communication: allowing language models to be fine-tuned to meet audience needs will allow for health education materials to be rapidly adapted with high-quality training materials and output review by a domain expert. For chatbots connected to EMR (Electronic Medical Record) systems, patient communication styles and needs could be updated based on patient records. ChatGPT offers a significant opportunity for producing evidence-based KT tools for education that are rapidly adaptable to meet their health literacy and other needs. Finally, chatbots may be used in virtual patient creation, enabling better simulation of diverse patients with a definable core set of similar characteristics (e.g., ibuprofen is equally effective and in the same situations), and variability among others.

## Data Availability

All data produced in the present study are available upon reasonable request to the authors

## Author contribution

JB had full access to all data in the study and takes responsibility for the integrity of the data and the accuracy of data analysis.

## Data availability

All data produced in the present study are available upon reasonable request to the author.

## Acknowledgements

I would like to thank my postdoctoral fellowship supervisors Dr. Shannon Scott and Dr. Lisa Hartling for their expert review and input into the draft manuscript.

This work was supported in part by Oracle Cloud credits and related resources provided by the Oracle for Research program.

## References

1. Gourlay A, Mshana G, Birdthistle I, Bulugu G, Zaba B, Urassa M. 2014. Using vignettes in qualitative research to explore barriers and facilitating factors to the uptake of prevention of mother-to-child transmission services in rural tanzania: A critical analysis. BMC Med Res Methodol. 14:21.

2. Stacey D, Briere N, Robitaille H, Fraser K, Desroches S, Légaré F. 2014. A systematic process for creating and appraising clinical vignettes to illustrate interprofessional shared decision making. Journal of Interprofessional Care. 28(5):453–459.

3. Murphy J, Hughes J, Read S, Ashby S. 2022. Evidence and practice: A review of vignettes in qualitative research. Nurse Researcher. 30(2).

4. Royal KD, Hedgpeth M-W, Jeon T, Colford CM. 2018. Automated item generation: The future of medical education assessment? INNOVATIONS.

5. St. Marie B, Jimmerson A, Perkhounkova Y, Herr K. 2021. Developing and establishing content validity of vignettes for health care education and research. Western Journal of Nursing Research. 43(7):677–685.

6. Cheng C, Elsworth GR, Osborne RH. 2020. Co-designing ehealth and equity solutions: Application of the ophelia (optimizing health literacy and access) process. Frontiers in public health. 8:604401.

7. Tremblay D, Turcotte A, Touati N, Poder TG, Kilpatrick K, Bilodeau K, Roy M, Richard PO, Lessard S, Giordano É. 2022. Development and use of research vignettes to collect qualitative data from healthcare professionals: A scoping review. BMJ Open. 12(1):e057095.

8. Matic R, Kabiljo M, Zivkovic M, Cabarkapa M. 2021. Extensible chatbot architecture using metamodels of natural language understanding. Electronics. 10(18):2300.

9. An overview of chatbot technology. Artificial Intelligence Applications and Innovations: 16th IFIP WG 125 International Conference, AIAI 2020, Neos Marmaras, Greece, June 5–7, 2020, Proceedings, Part II 16; 2020: Springer.

10. Jungmann SM, Klan T, Kuhn S, Jungmann F. 2019. Accuracy of a chatbot (ada) in the diagnosis of mental disorders: Comparative case study with lay and expert users. JMIR Formative Research. 3(4).

11. Luo B, Lau RYK, Li C, Si Y-W. 2022. A critical review of state-of-the-art chatbot designs and applications. WIREs Data Mining and Knowledge Discovery. 12(1):e1434.

12. Gilson A, Safranek C, Huang T, Socrates V, Chi L, Taylor RA, Chartash D. 2022. How well does chatgpt do when taking the medical licensing exams? The implications of large language models for medical education and knowledge assessment. medRxiv.2022.2012. 2023.22283901.

13. Chen M, Tworek J, Jun H, Yuan Q, Pinto HPdO, Kaplan J, Edwards H, Burda Y, Joseph N, Brockman G. 2021. Evaluating large language models trained on code. arXiv preprint arXiv:210703374.

14. Borsos Z, Marinier R, Vincent D, Kharitonov E, Pietquin O, Sharifi M, Teboul O, Grangier D, Tagliasacchi M, Zeghidour N. 2022. Audiolm: A language modeling approach to audio generation. arXiv preprint arXiv:220903143.

15. Daras G, Dimakis AG. 2022. Discovering the hidden vocabulary of dalle-2. arXiv preprint arXiv:220600169.

16. Poole B, Jain A, Barron JT, Mildenhall B. 2022. Dreamfusion: Text-to-3d using 2d diffusion. arXiv preprint arXiv:220914988.

17. Villegas R, Babaeizadeh M, Kindermans P-J, Moraldo H, Zhang H, Saffar MT, Castro S, Kunze J, Erhan D. 2022. Phenaki: Variable length video generation from open domain textual description. arXiv preprint arXiv:221002399.

18. Alayrac J-B, Donahue J, Luc P, Miech A, Barr I, Hasson Y, Lenc K, Mensch A, Millican K, Reynolds M. 2022. Flamingo: A visual language model for few-shot learning. arXiv preprint arXiv:220414198.

19. Gozalo-Brizuela R, Garrido-Merchan EC. 2023. Chatgpt is not all you need. A state of the art review of large generative ai models. arXiv preprint arXiv:230104655.

20. Openai. 2023. [accessed 2023 Jan 25]. https://openai.com/.

21. Semigran HL, Linder JA, Gidengil C, Mehrotra A. 2015. Evaluation of symptom checkers for self diagnosis and triage: Audit study. bmj. 351.

22. Chatgpt. 2023. [accessed]. https://chat.openai.com/chat.

23. Campbell A, Hartling L, Louie-Poon S, Scott SD. 2019. Parents’ information needs and preferences related to bronchiolitis: A qualitative study. CMAJ Open. 7(4):E640–e645.

24. Thompson AP, Nesari M, Hartling L, Scott SD. 2020. Parents’ experiences and information needs related to childhood fever: A systematic review. Patient Educ Couns. 103(4):750–763.

25. Archibald MM, Scott SD. 2019. Learning from usability testing of an arts-based knowledge translation tool for parents of a child with asthma. Nurs Open. 6(4):1615–1625.

26. Meherali S, Campbell A, Hartling L, Scott S. 2019. Understanding parents’ experiences and information needs on pediatric acute otitis media: A qualitative study. J Patient Exp. 6(1):53–61.

27. Reid K, Hartling L, Ali S, Le A, Norris A, Scott SD. 2017. Development and usability evaluation of an art and narrative-based knowledge translation tool for parents with a child with pediatric chronic pain: Multi-method study. J Med Internet Res. 19(12):e412.

28. Scott SD, Albrecht L, Given LM, Hartling L, Johnson DW, Jabbour M, Klassen TP. 2018. Pediatric information seeking behaviour, information needs, and information preferences of health care professionals in general emergency departments: Results from the translating emergency knowledge for kids (trekk) needs assessment. Cjem. 20(1):89–99.

29. Scott SD, Brett-MacLean P, Archibald M, Hartling L. 2013. Protocol for a systematic review of the use of narrative storytelling and visual-arts-based approaches as knowledge translation tools in healthcare. Syst Rev. 2:19.

30. Scott SD, Hartling L, O’Leary KA, Archibald M, Klassen TP. 2012. Stories – a novel approach to transfer complex health information to parents: A qualitative study. Arts & Health. 4(2):162–173.

31. Hartling L, Scott S, Pandya R, Johnson D, Bishop T, Klassen TP. 2010. Storytelling as a communication tool for health consumers: Development of an intervention for parents of children with croup. Stories to communicate health information. BMC Pediatr. 10:64.

32. Hartling L, Scott SD, Johnson DW, Bishop T, Klassen TP. 2013. A randomized controlled trial of storytelling as a communication tool. PLoS One. 8(10):e77800.

33. Li MH, Yang YS, Wang KC, Chiang YH, Shen PS, Lai WC, Guo TF, Chen P. 2017. Robust and recyclable substrate template with an ultrathin nanoporous counter electrode for organic-hole-conductor-free monolithic perovskite solar cells. ACS Appl Mater Interfaces. 9(48):41845–41854.

34. Ikah DS, Finn GM, Swamy M, White PM, McLachlan JC. 2015. Clinical vignettes improve performance in anatomy practical assessment. Anatomical sciences education. 8(3):221–229.

35. Rousseau A, Rozenberg P, Ravaud P. 2015. Assessing complex emergency management with clinical case-vignettes: A validation study. PLoS One. 10(9):e0138663.

36. Russo PL, Barnett AG, Cheng AC, Richards M, Graves N, Hall L. 2015. Differences in identifying healthcare associated infections using clinical vignettes and the influence of respondent characteristics: A cross-sectional survey of australian infection prevention staff. Antimicrobial Resistance and Infection Control. 4(1):1–7.

37. Haider AH, Schneider EB, Sriram N, Scott VK, Swoboda SM, Zogg CK, Dhiman N, Haut ER, Efron DT, Pronovost PJ. 2015. Unconscious race and class biases among registered nurses: Vignette-based study using implicit association testing. Journal of the American College of Surgeons. 220(6):1077-1086. e1073.

38. Ben-Shabat N, Sharvit G, Meimis B, Joya DB, Sloma A, Kiderman D, Shabat A, Tsur AM, Watad A, Amital H. 2022. Assessing data gathering of chatbot based symptom checkers-a clinical vignettes study. International Journal of Medical Informatics. 168:104897.

39. Hammoud M, Douglas S, Darmach M, Alawneh S, Sanyal S, Kanbour Y. 2022. Avey: An accurate ai algorithm for self-diagnosis. medRxiv.2022.2003. 2008.22272076.

40. Ma C. 2020. Vignette element analysis for automated generation of vignettes in pharmacy education.

41. Gao CA, Howard FM, Markov NS, Dyer EC, Ramesh S, Luo Y, Pearson AT. 2022. Comparing scientific abstracts generated by chatgpt to original abstracts using an artificial intelligence output detector, plagiarism detector, and blinded human reviewers. bioRxiv.2022.2012. 2023.521610.

42. Jeblick K, Schachtner B, Dexl J, Mittermeier A, Stüber AT, Topalis J, Weber T, Wesp P, Sabel B, Ricke J. 2022. Chatgpt makes medicine easy to swallow: An exploratory case study on simplified radiology reports. arXiv preprint arXiv:221214882.

43. Semigran HL, Levine DM, Nundy S, Mehrotra A. 2016. Comparison of physician and computer diagnostic accuracy. JAMA Intern Med. 176(12):1860–1861.

44. Kung TH, Cheatham M, Medinilla A, ChatGPT, Sillos C, De Leon L, Elepano C, Madriaga M, Aggabao R, Diaz-Candido G. 2022. Performance of chatgpt on usmle: Potential for ai-assisted medical education using large language models. medRxiv.2022.2012. 2019.22283643.

45. Ceney A, Tolond S, Glowinski A, Marks B, Swift S, Palser T. 2021. Accuracy of online symptom checkers and the potential impact on service utilisation. PLOS ONE. 16(7):e0254088.

46. Wallace W, Chan C, Chidambaram S, Hanna L, Iqbal FM, Acharya A, Normahani P, Ashrafian H, Markar SR, Sounderajah V et al. 2021. The diagnostic and triage accuracy of digital and online symptom checker tools: A systematic review. medRxiv.2021.2012.2021.21268167.

